# Genomic epidemiology reveals the impact of national and international restrictions measures on the SARS-CoV-2 epidemic in Brazil

**DOI:** 10.1101/2021.10.07.21264644

**Authors:** Marta Giovanetti, Svetoslav Nanev Slavov, Vagner Fonseca, Eduan Wilkinson, Houriiyah Tegally, José Salvatore Leister Patané, Vincent Louis Viala, James Emmanuel San, Evandra Strazza Rodrigues, Elaine Vieira Santos, Flavia Aburjaile, Joilson Xavier, Hegger Fritsch, Talita Emile Ribeiro Adelino, Felicidade Pereira, Arabela Leal, Felipe Campos de Melo Iani, Glauco de Carvalho Pereira, Cynthia Vazquez, Gladys Mercedes Estigarribia Sanabria, Elaine Cristina de Oliveira, Luiz Demarchi, Julio Croda, Rafael dos Santos Bezerra, Loyze Paola Oliveira de Lima, Antonio Jorge Martins, Claudia Renata dos Santos Barros, Elaine Cristina Marqueze, Jardelina de Souza Todao Bernardino, Debora Botequio Moretti, Ricardo Augusto Brassaloti, Raquel de Lello Rocha Campos Cassano, Pilar Drummond Sampaio Corrêa Mariani, João Paulo Kitajima, Bibiana Santos, Rodrigo Proto-Siqueira, Vlademir Vicente Cantarelli, Stephane Tosta, Vanessa Brandão Nardy, Luciana Reboredo de Oliveira da Silva, Marcela Kelly Astete Gómez, Jaqueline Gomes Lima, Adriana Aparecida Ribeiro, Natália Rocha Guimarães, Luiz Takao Watanabe, Luana Barbosa Da Silva, Raquel da Silva Ferreira, Mara Patricia F. da Penha, María José Ortega, Andrea Gómez de la Fuente, Shirley Villalba, Juan Torales, María Liz Gamarra, Carolina Aquino, Gloria Patricia Martínez Figueredo, Wellington Santos Fava, Ana Rita C. Motta-Castro, James Venturini, Sandra Maria do Vale Leone de Oliveira, Crhistinne Cavalheiro Maymone Gonçalves, Maria do Carmo Debur Rossa, Guilherme Nardi Becker, Mayra Marinho Presibella, Nelson Quallio Marques, Irina Nastassja Riediger, Sonia Raboni, Gabriela Mattoso Coelho, Allan Henrique Depieri Cataneo, Camila Zanluca, Claudia N Duarte dos Santos, Patricia Akemi Assato, Felipe Allan da Silva da Costa, Mirele Daiana Poleti, Jessika Cristina Chagas Lesbon, Elisangela Chicaroni Mattos, Cecilia Artico Banho, Lívia Sacchetto, Marília Mazzi Moraes, Rejane Maria Tommasini Grotto, Jayme A. Souza-Neto, Maurício Lacerda Nogueira, Heidge Fukumasu, Luiz Lehmann Coutinho, Rodrigo Tocantins Calado, Raul Machado Neto, Ana Maria Bispo de Filippis, Rivaldo Venancio da Cunha, Carla Freitas, Cassio Roberto Leonel Peterka, Cássia de Fátima Rangel Fernandes, Wildo Navegantes de Araújo, Rodrigo Fabiano do Carmo Said, Maria Almiron, Carlos Frederico Campelo de Albuquerque e Melo, José Lourenço, Tulio de Oliveira, Edward C. Holmes, Ricardo Haddad, Sandra Coccuzzo Sampaio, Maria Carolina Elias, Simone Kashima, Luiz Carlos Junior de Alcantara, Dimas Tadeu Covas

## Abstract

Brazil has experienced some of the highest numbers of COVID-19 cases and deaths globally and from May 2021 made Latin America a pandemic epicenter. Although SARS-CoV-2 established sustained transmission in Brazil early in the pandemic, important gaps remain in our understanding of virus transmission dynamics at the national scale. Here, we describe the genomic epidemiology of SARS-CoV-2 using near-full genomes sampled from 27 Brazilian states and a bordering country - Paraguay. We show that the early stage of the pandemic in Brazil was characterised by the co-circulation of multiple viral lineages, linked to multiple importations predominantly from Europe, and subsequently characterized by large local transmission clusters. As the epidemic progressed under an absence of effective restriction measures, there was a local emergence and onward international spread of Variants of Concern (VOC) and Variants Under Monitoring (VUM), including Gamma (P.1) and Zeta (P.2). In addition, we provide a preliminary genomic overview of the epidemic in Paraguay, showing evidence of importation from Brazil. These data reinforce the usefulness and need for the implementation of widespread genomic surveillance in South America as a toolkit for pandemic monitoring that provides a means to follow the real-time spread of emerging SARS-CoV-2 variants with possible implications for public health and immunization strategies.

## Introduction

At the end of 2019, a novel respiratory pathogen designated the Severe Acute Respiratory Syndrome Coronavirus 2 (SARS-CoV-2) emerged in the city of Wuhan, China. Since its identification, the virus has spread rapidly on a global scale causing an unprecedented Coronavirus disease 2019 (COVID-19) pandemic (declared on March 11^th^ 2020) which has overwhelmed many health care systems^1^. By the mid-February 2022, more than 415 million cases of COVID-19, with more than 5.84 million associated deaths, have been reported globally^2^. Brazil has become the epicentre of the COVID-19 epidemic in the Americas, with a total of 21.2 million cases and a death toll exceeding 591,000 reported cases by the end of September of 2021, making it one of the countries hardest hit by COVID-19^3^. Critically, however, a lack of genome sequence data from Brazil has limited our ability to fully understand transmission dynamics at the national scale.

To assess how population restriction measures might have played a role in shaping viral diversity and evolution during the SARS-CoV-2 epidemic in Brazil, we performed a phylogenetic and phylogeographic analysis of genomic data from 27 Brazilian states and one neighbouring country – Paraguay – collected up to September 2021. We show that the early spread of SARS-CoV-2 in Brazil resulted from initial founder events associated with international travel (mainly from Europe), which then spread through regional mobility pathways during the recession of the first epidemic wave. From this, Brazil emerged as an exporter of SARS-CoV-2 variants to other countries. We further describe the emergence and spread of key viral Variants of Concern (VOCs; for example, Gamma) and Variants Under Monitoring (VUMs; for example, Zeta) in Brazil, highlighting how their emergence may have contributed to a more severe second wave in the country.

## Results

### COVID-19 transmission dynamics in Brazil

The first confirmed infection of SARS-CoV-2 in Brazil was on February 26^th^ 2020 in the State of São Paulo (SP), in a traveller returning from Italy (**Fig. 1A**). On March 17^th^ 2020 the first COVID-19-related death, a 61-year old male, was reported in the same state^4,5^. Four days later, all Brazilian states reported at least one confirmed case of COVID-19 and the Brazilian Ministry of Health (BR-MoH) declared an outbreak of large-scale community transmission of the virus^6^. By April 10^th^, 2020 the virus had already reached remote locations such as the Yanomami indigenous community located in the state of Roraima, in northern Brazil^6^ (**Fig. 1A**).

**Fig. 1.**
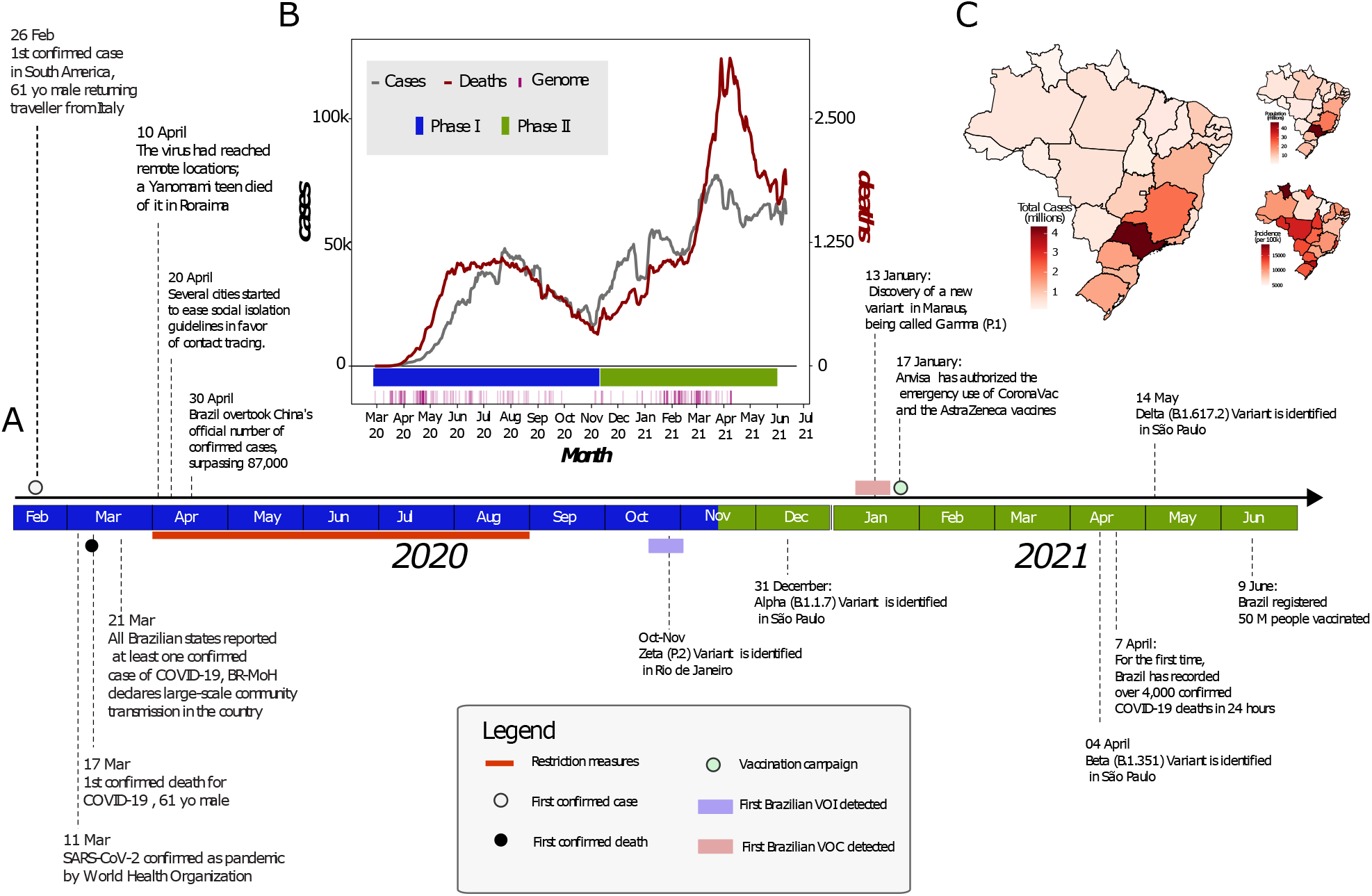
Key events following the first confirmed infection of SARS-CoV-2 in Brazil. (A) Timeline of SARS-CoV-2 key events in Brazil. The Brazilian map was colored according to geographical macro region: North (red), Northeast (green), Southeast (purple), Midwest (light blue) South (light orange). (B) Epidemic curve showing the progression of reported daily viral infection numbers in Brazil from the beginning of the epidemic (grey) and deaths (red) in the same period, with restriction phases indicated along the bottom. (C) Map of cumulative SARS-CoV-2 cases per 100,000 inhabitants in Brazil up to June 2021.

After the World Health Organization (WHO) declared the outbreak of SARS-CoV-2 as a public health emergency of international concern on January 30^th^ 2020 the Brazilian government introduced restriction measures to mitigate viral spread (**Fig. 1A**)^7^. The primary measure involved social isolation, followed by the closure of schools, universities, and non-essential businesses^8^. Additional measures included the mandatory use of personal protective masks^9^, the cancellation of events expected to attract large numbers of people and tourists, and only services considered as essential such as markets and pharmacies remained open^8,10^. However, while the epidemic was growing, restriction measures were progressively eased to mitigate negative impacts on the economy. Notably, even during periods of restriction, travel between Brazilian states largely remained possible, facilitating SARS-CoV-2 transmission throughout the country^11^. The latter was likely linked to the emergence of new and more contagious viral lineages documented as VOC - Gamma (lineage P.1) - and VUM - Zeta (lineage P.2), which may also have contributed to a more severe second wave (**Fig. 1B**)^11,13,14,15^.

The COVID-19 death toll in Brazil steadily rose from March 2021. It reached a daily total of 4,250 on April 2021, the highest number of daily fatalities from COVID-19 worldwide (**Fig. 1B**). Signs of collapse of the health system were reported in numerous cities among different regions of the country. The situation worsened considerably following the emergence of multiple VOCs and VUMs in parallel with a slow progress of the vaccination campaign^16^. Vaccination in Brazil began on January 17^th^ 2021 when the Instituto Butantan imported the first 6 million doses of CoronaVac (a whole virus inactivated vaccine) in a collaboration with Sinovac Biotech (**Fig. 1A**)^17,18^. As of February 16^th^ 2022 approximately 71.8% of the Brazilian population have been vaccinated with the first dose of any of the vaccines currently available (CoronaVac, AstraZeneca, Pfizer and Janssen), and only 22% had been fully vaccinated (i.e. a single dose of Janssen or both doses of the other vaccines)^19^.

By analysing the total number of COVID-19 notified cases to the end of September 2021, we observed that the Brazilian region with the highest population density (Southeast) also contained the highest number of the cases registered in the country, with the state of São Paulo documenting the largest number of cases (n=4,369,410) in that period (**Fig. 1C**). However, when we consider the incidence rate (number of reported cases/population) by state, we found that the Midwest, the least populated region in Brazil, had the highest incidence rate, with 13,604.23 cases/100K inhabitants^1^.

### SARS-CoV-2 genomic data

A total of 3,866 near-full genomes sequences from SARS-CoV-2 RT-qPCR positive samples were obtained as part of this study. SARS-CoV-2 sequencing spanned February 2020 to June 2021, sampled from 8 of the 27 Brazilian states (São Paulo=3309; Rio Grande do Sul=48; Paraná=55; Minas Gerais=80; Mato Grosso do Sul=36; Mato Grosso=51; Bahia=224) and one neighbouring country, Paraguay (n=63). Almost half of the sequences were from Southeast Brazil, consistent with it reporting most cases natiionally (**Fig. 1C**)^6^. The sequenced samples were collected from 2,023 females and 1,843 males (**Table S1 and S2**) with a median age of 41.72 years (range: 1 to 90 years of age). All tested samples contained sufficient viral genetic material (≥2ng/µL) for library preparation. For positive samples, PCR cycle threshold (Ct) values were on average 19.93 (range: 10.75 to 30). Sequences had a median genome coverage of 95% (range: 80 to 99.99) and average genome coverage was typically higher for samples with lower Ct values (**Fig. S1**). Epidemiological information and sequencing statistics of the generated sequences from Brazil and Paraguay are detailed in **Tables S1** and **S2**, respectively. Sequences were assigned to 39 different PANGO-lineages based on the proposed dynamic nomenclature for SARS-CoV-2 lineages (**Fig. S1, Table S1 and S2**) and have been submitted to GISAID following the WHO guidelines (**Tables S1** and **S2**) (Pangolin version 3.1.7, August 2021).

### Phylogenetic inference and lineage diversity

The rapid spread of SARS-CoV-2, together with the reported circulation of several VOCs and VUMs in Brazil prompted the intensification of genomic surveillance by the National Network for Pandemic Alert of SARS-CoV-2 at the end of December 2020. As of June 30^th^ 2021 some 17,135 SARS-CoV-2 genomes from Brazil had been deposited in the GISAID database from all 27 Brazilian states (**Fig. 2A**). The states with the highest number of sequenced genomes were São Paulo (n=9,600) and Rio de Janeiro (n=2,031). Although genomic surveillance began as soon as the first confirmed infections were detected in Brazil, by the end of June 2021 there was still a paucity of genomic data from some states such as Roraima (n=29), Acre (n=29), Rondônia (n=37), Tocantins (n=27), Piauí (n=19) and the Federal District (n=33) (**Fig. 2A**). Half of all Brazilian genomes were deposited in early 2021, suggesting intensified surveillance in the second wave following the detection of Gamma (and other VOCs (e.g. Alpha/B.1.1.7) and VUMs (e.g. Zeta) throughout the country (**Fig. 2B**).

**Fig. 2.**
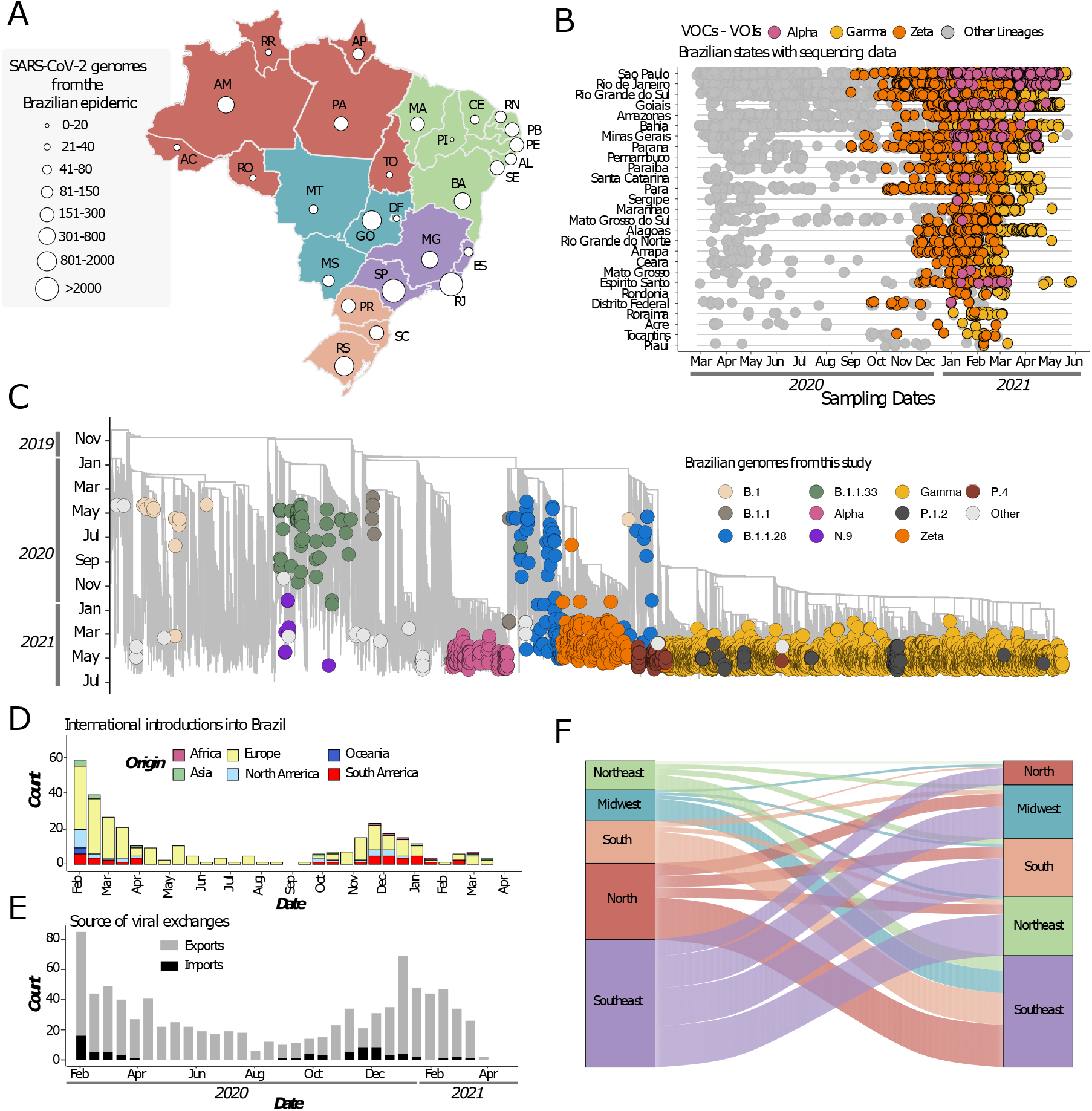
Phylogenetic analysis and SARS-CoV-2 lineage dynamics in Brazil. (A) Map of Brazil with the number of sequences in GISAID as of 30th June 2021. (B) Temporal sampling of sequences in Brazilian states through time with VOCs highlighted and annotated according to their PANGO lineage assignment. (C) Time resolved maximum likelihood phylogeny containing high qualitynear-full-genome sequences from Brazil (n=3866) obtained from this study, analysed against a backdrop of global reference sequences (n=25,288). Variants under monitoring (VUM) and concern (VOC) are highlighted on the phylogeny. (D) Sources of viral introductions into Brazil characterized as external introductions from the rest of the world. (E) Sources of viral exchanges (imports and exports) in and outside Brazil. (F) Number of viral exchanges within Brazilian regions by counting the state changes from the root to the tips of the phylogeny in panel C.

To obtain a better understanding of the dynamics of SARS-CoV-2 spread in Brazil, we coupled epidemiological data with phylodynamic analysis for a data set comprising 25,288 available globally representative genomes, including the new genomes obtained in this study (n=3,866) sampled from December 26^th^, 2019 to June 28^th^ 2021 (**Fig. 2C, Fig. S2**). A date-stamped phylogeny of these data indicated that most of the Brazilian sequences were interspersed with those introduced from several countries (**Fig. 2C, 2D**). This pattern further indicated that the co-circulation of multiple SARS-CoV-2 lineages over time was linked to multiple importations followed by large local transmissions concomitant with a high number of infections (**Fig. 2C, 2D**).

Using an ancestral location state reconstruction on the dated phylogeny we were able to infer the number of viral imports and exports between Brazil and the rest of the world, and between individual Brazilian regions (hereafter referred as the North, Northeast, Midwest, Southeast and South regions) (**Fig. 2D-F**). The bulk of imported introductions (estimated to be 114 independent ones) were largely from Europe (**Fig. 2D**), occurring before the implementation of restriction measures (April 2020) when the epidemic was rapidly progressing (**Fig. 2D, E**). However, at least 33 introduction events are inferred to have occurred during enforcement of preventive measures up to August 2020 (**Fig. 2D, E**), and hence before those measures were loosened. Finally, although Brazil was a major virus importer, there were approximately 10 times more inferred exportation events out of Brazil than viral introductions into Brazil (**Fig. 2E**).

Our estimates of viral movement within Brazil further suggested that the Southeast region was the largest contributor of viral exchanges to other regions, comprising approximately 40% of viral movements from one geographical region to another, followed by the North region that contributed to approximately 25% of all viral movements. Although these estimates are in line with epidemiological data, this observation is also likely to be influenced by these two regions having the greatest number of sequences available for analysis.

### Spatiotemporal spreading of Brazilian VOC (Gamma) and VUM (Zeta)

We next focused on the two Brazilian variants that evolved from the B.1.1.28 lineage and grew into large transmission clusters during the second wave of the epidemic from January 2021 -the VOC (Gamma/P.1) and VUM (Zeta/P.2). To assess the detailed evolution of these lineages over time we performed a spatiotemporal phylogeographic analysis using a molecular clock model.

The Gamma VOC was first sampled in Brazil in early January 2021^12,20^. It displayed an unusual number of lineage-defining mutations in the S protein, including three designated that may impact transmission, immune escape, and virulence - N501Y, E484K and K417T^21–23^. In line with previous estimates^12,20^, our phylogeographic analysis suggested that the Gamma variant emerged around November 21^st^ 2020 (95% highest posterior density, 12-29 November 2020) in Manaus (Amazonas state) in Northern Brazil and spread extensively among Brazilian regions (**Fig. 3A, C**). Our data reveals multiple introductions of this lineage from the Amazonas state to Brazil’s southeastern, northeastern and midwestern states (**Fig. 3A, C**). By mid-January 2021, the southeastern and northern regions had also acted as source populations for the introduction of this variant into the southern region (**Fig 3A, C**).

**Fig. 3.**
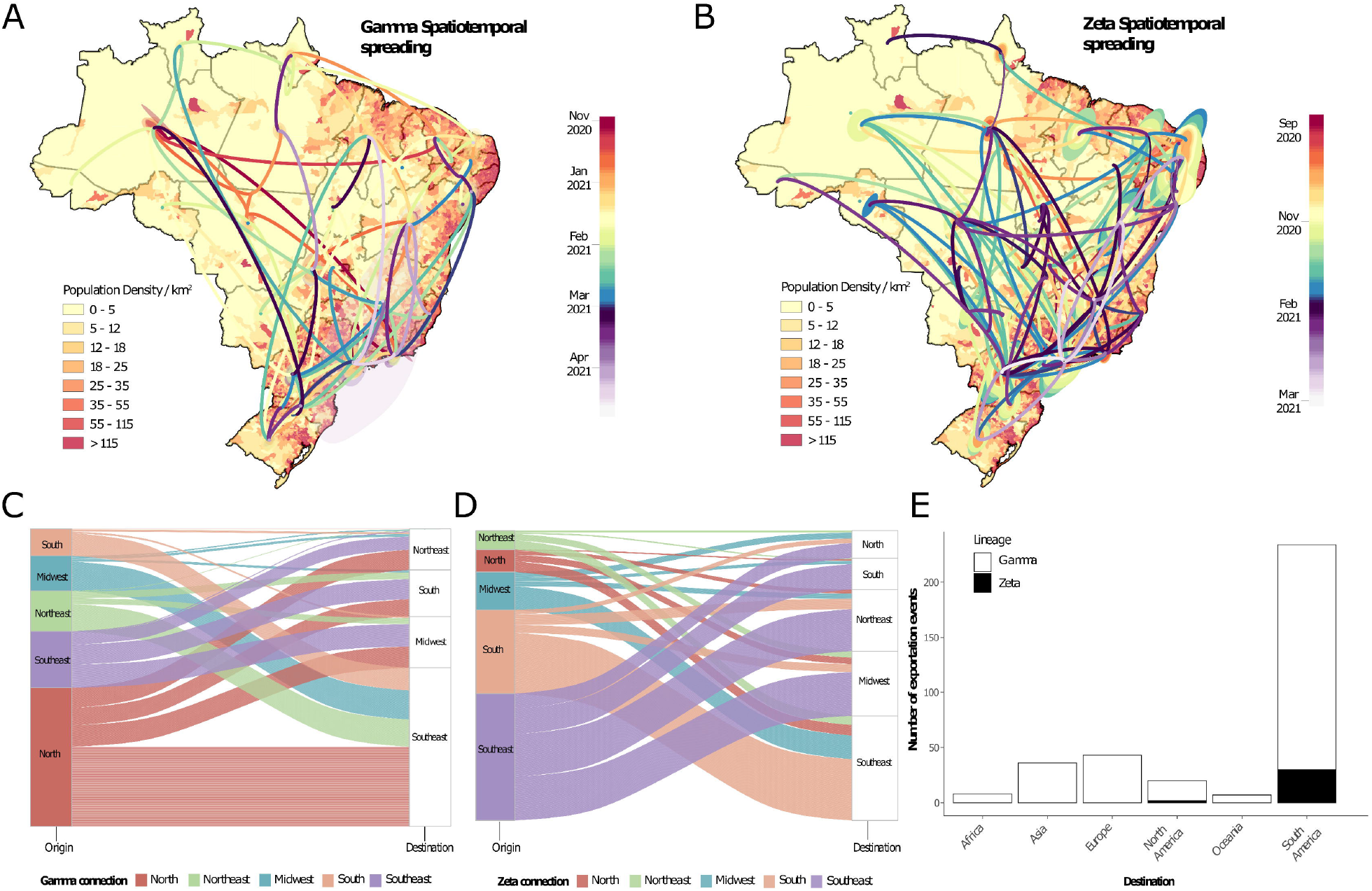
Spatiotemporal spread of VOC and VUM in Brazil. (A) Phylogeographic reconstruction of the spread of the Gamma VOC in Brazil. Circles represent nodes of the maximum clade credibility phylogeny and are colored according to their inferred time of occurrence. Shaded areas represent the 80% highest posterior density interval and depict the uncertainty of the phylogeographic estimates for each node. Differences in population density are shown on a dark-white scale; B) Phylogeographic reconstruction of the spread of the Zeta VUM across Brazil. Circles represent nodes of the maximum clade credibility phylogeny and are colored according to their inferred time of occurrence. Shaded areas represent the 80% highest posterior density interval and depict the uncertainty of the phylogeographic estimates for each node. Differences in population density are shown on a dark-white scale; In both Panels (A) and (B) solid curved lines denote the links between nodes and the directionality of movement is anti-clockwise along the curve (as shown in the “dispersal direction” sublegends). (C) Number of exchanges of the Gamma variant between Brazilian regions (N=North; NE=Northeast; MD=Midwest; SE=Southeast; S=South); (D) Number of exchanges of the Zeta variant between Brazilian regions; (E) Sources of viral export of the VOC and VUM from Brazil to the rest of the world.

Zeta (P.2) is defined by the presence of the S:E484K mutation in the receptor binding domain (RBD) and other lineage-defining mutations outside the S protein^15,24^. Although it was first described in samples from October 2020 in the state of Rio de Janeiro, our phylogeographic reconstruction suggests that the variant originated in Paraná state in South Brazil late August 2020 (95% highest posterior density, 19 August 2020 to 03 September 2020) (**Fig. 4B**). Since then, Zeta has spread multiple times to much of the southeastern, northeastern, midwestern and northern Brazilian regions (**Fig. 4D**). Together, our results further suggest that the transmission dynamics roughly followed patterns of population density, moving most often between the most populous localities (**Fig. A, B**).

**Fig. 4.**
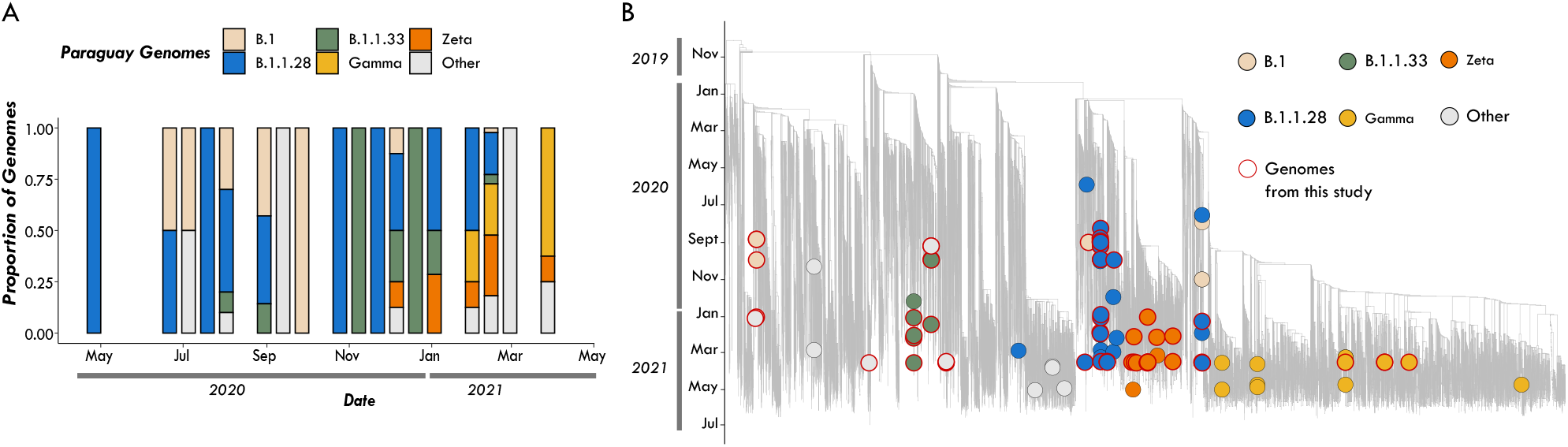
The SARS-CoV-2 epidemic in Paraguay. (A) Epidemic curve showing the progression of reported viral infection numbers in Paraguay from the beginning of the epidemic (grey) and deaths (red) in the same period; (B) Progressive distribution of the top 20 PANGO lineages in Paraguay over time; (C) Time resolved maximum likelihood tree containing (n=63) high quality near-complete genome sequences from Paraguay obtained in this study analysed against a backdrop of global reference sequences. VUM and VOCs are highlighted on the phylogeny. Small circles indicate genome sequences from Brazil. Bigger circles indicate sequences from Paraguay. New genomic sequences from Paraguay obtained in this study are highlighted with a red border.

By estimating the pattern of migration flows we also examined the potential role of Brazil as an exporter of the Gamma and Zeta variants to the rest of the world (**Fig. 3**). While the North region seeded approximately 47% of all Gamma infections into other regions, consistent with it being where this lineage originated, there is strong evidence both from phylogeographic analysis (**Fig. 3A**) and ancestral state reconstruction (**Fig. 3C**) that there was considerable subsequent transfer of Gamma between all regions. Zeta had a different dispersal pattern to Gamma, with 73% of all Zeta movements originating in the Southeast and South regions, consistent with our phylogeographic reconstruction that this is the geographic source of this lineage (**Fig. 3**).

Our analysis further revealed that Brazil has contributed to the international spread of both variants with at least 316 and 32 exportation events to the rest of the world detected for Gamma and Zeta variants, respectively (**Fig. 3E**). Consistent with importations, most exports were to South America (65%) and Europe (14%), followed by Asia (11%), North America (5%), Africa (2.5%) and Oceania (2.5%) with an increase between January 2021 and March 2021 coinciding with the second wave of infections in Brazil and some relaxation of international travel restrictions (**Fig. 3E**). As shown elsewhere, these results demonstrate that under relaxation of travel restrictions SARS-CoV-2 lineages can spread to a diverse range of international locations^25-30^.

### Cross-border SARS-CoV-2 transmission from Brazil to Paraguay

To explore the burden of the Brazilian SARS-CoV-2 pandemic on other South American countries, we provide a preliminary overview of the SARS-CoV-2 epidemic in Paraguay. The first COVID-19 confirmed case was documented in Paraguay on March 7^th^ 2020 in a 32-year-old man from San Lorenzo, Central Department. Thirteen days later, the first death and the first case of community transmission were also confirmed. COVID-19 cases in Paraguay rose sharply in March (**Fig. 4A**) resulting in 100% occupancy of intensive care beds, prompting the government to declare a strict quarantine to mitigate the spread of the virus^31,32^. By the end of June 2021 a total number of 460,000 confirmed cases and 15,000 coronavirus-related deaths had been reported in Paraguay^31^.

The COVID-19 epidemic in Paraguay can generally be characterized by three phases: phase I staring from March 10^th^ 2020 characterized by restriction measures; phase II since May 4^th^, 2020, also called “Intelligent/Smart Quarantine” with a gradual return to work and social activities; and phase III implemented since October 5^th^ 2020, known as the “COVID way of living”, characterized by the relaxation of the restrictions measures and the reopening of national borders and resumption of international flights^32^.

Since the beginning of the epidemic there has been a paucity of whole genomes sequences from Paraguay, with only n=165 whole genome sequences available on GISAID by the end of July 2021, about 0.0003% of known cases. This seriously impacts the ability to characterize the molecular epidemiology of SARS-CoV-2 at a regional level. In collaboration with the Pan-American Health Organization and the National Public Health Laboratory of Asunción in Paraguay we obtained a total of 63 near complete genome sequences sampled between July 2020 and June 2021, representing ∼ 40% of the currently available genomes from this country. The selection of the samples was based on the Ct value (≤30) and availability of epidemiological metadata, such as date of sample collection, sex, age and municipality of residence. Thus, by applying these inclusion criteria only 63 positive samples were considered suitable for this study. As expected, we observed the co-circulation of multiple SARS-CoV-2 lineages **(Fig. 4B**), linked to multiple importations and subsequently characterized by large transmission clusters.

Importantly, our phylogenetic analysis revealed that most of the SARS-CoV-2 variants currently circulating in Paraguay, including lineages (B.1.1.28; B.1.1.33; Zeta and Gamma), originally emerged in Brazil (Fig. 4A and Fig.4B), thus suggesting cross-border transmission from Brazil to Paraguay (**Fig. 4B, C**). This reinforces the importance of nonpharmaceutical measures in containing and preventing the spread of viral strains into neighbouring countries.

As of July 31^th^, 2021 a total of 78% of available genomic sequences from Paraguay were linked to infections caused by Brazilian variants, with the Gamma VOC being the most prevalent lineage in the country. As genome sequencing is not widespread, it is difficult to determine how widely these variants have spread within Paraguay and to other Latin American countries. However, the abundant COVID-19 cases in Brazil, a country that shares borders with ten countries, suggests that this risk is likely to be high.

## Discussion

Genome sequencing combined with epidemiological assessment has been widely used as a tool to track the spread and evolution of SARS-CoV-2, enabling public health authorities to tailor their control strategies. In addition, genomic data can reveal key properties of emerging viruses, including precise epidemiological behaviour that patient data alone cannot capture. However, the utility of genomic data is marred by sparse sampling and geographic inequities^33^. Brazil is an important case study, but SARS-CoV-2 sequences from Brazil are only available from a tiny fraction of the number of confirmed cases into the country, curbing their utility in a public health setting and limiting effective control strategies.

To help overcome these limitations, we report genomic data obtained by sequencing 3,866 SARS-CoV-2 infection cases confirmed by RT-qPCR from patients residing in 8 of the 27 Brazilian federal states and the neighbouring country of Paraguay. These were analysed together with n=13,328 and n=102 (up to 30, June 20^th^ 2021) publicly available complete genomes sequences from both countries, respectively. By combining epidemiological and genomic data, we show how the interplay between the implementation of restriction measures and sustained SARS-CoV-2 transmission have shaped the Brazilian epidemic over 20 months, including the dramatic resurgences in case numbers linked with the emergence of VOCs and VUMs. In particular, we show that multiple independent importations of SARS-CoV-2, predominantly from Europe, had occurred in Brazil during the early phase of the epidemic (up to April 2020). We further detected multiple (n=33) international introductions during periods characterized by the enforcement of preventive measures, demonstrating that these can be ineffective to prevent importation, as previously reported in other countries^34^.

Our analysis shows that viral importations were present throughout the entirety of the study period, except between May and September 2020 and further suggest that Brazil played much more as a viral exporter, resulting in 10 times more inferred exportations events than viral introduction into the country (**Fig. 2E**). Importantly, this was also a period linked to the emergence of Brazilian VOCs and VUMs. Although we have not measured passenger traffic to and from Brazil, the viral migration pattern observed appears to be in line with results obtained from other studies combining viral genetic data with epidemiological and travel data, and provide evidence that the dispersion of respiratory viruses is dictated by human mobility and population density^26-30^.

We also provide the first preliminary overview of the SARS-CoV-2 epidemic in Paraguay, revealing a high virus connectivity with Brazil. This spread was likely facilitated by airplane and road networks that form major transport trails linking Brazil to other parts of South America. The rapid unhampered spread of such VOCs and VUMs into South American countries, such as Paraguay, indicates that the land-border controls to curb the international spread of the virus have been largely ineffective and highlights the inherent challenges in screening cross-border travellers to contain the spread of the virus within this region. Our data further suggest that national and international travel restrictions lifted at certain points during the Brazilian epidemic were likely responsible for both virus introduction from international localities and within-country transmission, with infected travelers acting as carriers.

More broadly, our study highlights the utility of SARS-CoV-2 genome sequencing in COVID-19 outbreak investigation and the need for more comprehensive country-wide studies of the epidemiology and spread of emerging viral strains. Since the disproportion on the number of genome sequences available for each Brazilian state/region, our results further highlight the need of more public investments to strength the genomic capacity across the country.

Additionally, the current co-circulation of VUMs and VOCs in Brazil, together with the slow vaccine rollout, has important implications for public health in this high populous and regionally important country^35^. Such epidemiological conditions create the perfect environment for the continued evolution of SARS-CoV-2, potentially enabling the emergence of novel variants of altered phenotype.

## Materials and Methods

### Ethics statement

This research was approved by the Ethics Review Committee of the Pan American Health Organization (PAHOERC.0344.01), the Federal University of Minas Gerais (CEP/CAAE: 32912820.6.1001.5149), the University of São Paulo (CEP/FZEA: 4.780.992) and the Blood Center of Ribeirão Preto (CEP/HCRP-FMRP: 50367721.7.1001.5440), and by the Paraguayan Ministry of Public Health and Social Welfare (MSPyBS/ S.G. no. 0944/18). The availability of these samples for research purposes during outbreaks of national concern is allowed under the terms of the 510/2016 Resolution of the National Ethical Committee for Research – Brazilian Ministry of Health (CONEP - Comissão Nacional de Ética em Pesquisa, Ministério da Saúde) that authorizes, without the necessity of an informed consent, the use of clinical samples collected in the Brazilian Central Public Health Laboratories to accelerate knowledge building and contribute to surveillance and outbreak response. The samples processed in this study were obtained anonymously from material exceeding the routine diagnosis in Brazilian public health laboratories that belong to the public network within BrMoH.

### Genomic surveillance network

As a part of the National Network for Pandemic Alert of SARS-CoV-2, we performed genomic monitoring to rapidly understand the spread of SARS-CoV-2 at the national and cross border levels. This was performed in collaboration with the Central Laboratory of Public Health (LACEN) from the states of Bahia (BA), Mato Grosso (MT), Mato Grosso do Sul (MS), Minas Gerais (MG), as well as Blood Center of Ribeirão Preto, the Fiocruz Paraná (PR), the Butantan Institute from the state of São Paulo (SP) state, the University of São Paulo and the Central Laboratory of Health of Paraguay.

### Sample collection and molecular diagnostic assays

Convenience clinical samples from patients with suspected SARS-CoV-2 infection from patients residing in 8 of the 27 Brazilian federal states and from Asunción, Paraguay, and collected between July 2020 and June 2021 were provided for diagnostic and genome sequencing purposes. Viral RNA was extracted from nasopharyngeal swabs using an automated protocol and tested for SARS-CoV-2 by multiplex real-time PCR assays: (i) the Allplex 2019-nCoV Assay (Seegene) targeting the envelope (E), the RNA dependent RNA polymerase (RdRp) and the nucleocapsid (N) genes; (ii) the Charité: SARS-CoV2 (E/RP) assay (Bio-Manguinhos/Fiocruz) targeting the E gene, and (iii) the GeneFinder COVID-19 Plus RealAmp Kit (Osang Healthcare, South Korea) supplied by the BrMoH, Butantan Institute and the Pan-American Health Organization (OPAS).

### cDNA synthesis and whole genome sequencing

Samples were selected for sequencing based on the Ct value (≤30) and availability of epidemiological metadata, such as date of sample collection, sex, age and municipality of residence. The preparation of SARS-CoV-2 genomic libraries was performed using both the Illumina COVIDSeq test following the manufacturer’s instructions and nanopore sequencing using the ARTIC Network primal scheme (https://github.com/artic-network/artic-ncov2019/tree/master/primer_schemes/nCoV-2019/V3)^36^. The normalized libraries were loaded onto a 300-cycle MiSeq Reagent Kit v2 and run on the Illumina MiSeq instrument (Illumina, San Diego, CA, USA).

Due to regional characteristics and the accessibility of resources of the different Brazilian and Paraguayan laboratories, we also applied field SARS-CoV-2 sequencing using the Oxford Nanopore MinION technology. In this case the SuperScript IV Reverse Transcriptase kit (Invitrogen) was initially used for cDNA synthesis following the manufacturer’s instructions. The cDNA generated was subjected to multiplex PCR sequencing using the Q5 High Fidelity Hot-Start DNA Polymerase (New England Biolabs) and a set of specific primers designed by the ARTIC Network for sequencing the complete SARS-CoV-2 genome (Artic Network version 3)^36^. PCR conditions have been previously reported in^36^. All experiments were performed in a biosafety level-2 cabinet. Amplicons were purified using 1x AMPure XP Beads (Beckman Coulter) and quantified on a Qubit 3.0 fluorimeter (Thermofisher Scientific) using Qubit™ dsDNA HS Assay Kit (Thermofisher Scientific). DNA library preparation was performed using the Ligation Sequencing Kit LSK109(Oxford Nanopore Technologies) and the Native Barcoding Kit (NBD104 and NBD114, Oxford Nanopore Technologies). Sequencing libraries were loaded into a R9.4 flow cell (Oxford Nanopore Technologies). In each sequencing run, we used negative controls to prevent and check for possible contamination with less than 2% mean coverage.

### Generation of consensus sequences from Illumina and nanopore

The genome assembly pipeline for Illumina reads involved: (i) read trimming and filtering using Trimmomatic^37^; (ii) minimap2^38^for read mapping against the reference strain (Wuhan-Hu-1 genome reference - NCBI accession NC_045512.2); (iii) samtools^39^ for sorting and indexing; (iv) Pilon^40^ for improving the indel detection; (v) bwa mem^41^ for remapping against Pilon’s generated consensus; (vi) samtools mpileup to generate alignment quality values; (vii) seqtk^42^ to generate a quasi-final genome version; (viii) bwa mem for a 3rd round of remapping reads against the quasi-final genome; and (ix) samtools depth to assess position depths given the *.bam file from the previous step (nucleotide positions with read depth < 5 are denoted as “N”).

Oxford Nanopore sequencing raw files were basecalled using Guppy v3.4.5 and barcode demultiplexing was performed using qcat. Consensus sequences were generated by *de novo* assembling using Genome Detective^43^ that uses DIAMOND to identify and classify candidate viral reads in broad taxonomic units, using the viral subset of the Swissprot UniRef protein database. Candidate reads were next assigned to candidate reference sequences using NCBI blastn and aligned using AGA (Annotated Genome Aligner) and MAFFT. Final contigs and consensus sequences are then available as FASTA files.

### Data quality control and global data set collection

To ensure the quality of the genome sequences generated in this study and to guarantee the highest possible phylogenetic accuracy, only genomes >29,000bp and <1% of ambiguities were considered (n=3,866). Multiple sequence alignment was performed with MAFFT^44^ and a preliminary phylogenetic tree was inferred using the maximum likelihood method in IQ-TREE2, employing the GTR+I model of nucleotide substitution. Prior to further phylogenetic analysis our data set was also assessed for both sequences with low data quality (e.g. with assembling issues, sequencing and alignment errors, data annotation errors and sample contamination) and for molecular clock signal (i.e. temporal structure) using TempEst v1.5.3^45^.

We appended the 3,866 genome sequences newly generated under this project with an extensive reference data set of SARS-CoV-2 sequences sampled globally collected since the start of the outbreak, including n=13,328 near-complete genomes from Brazil and n=102 from Paraguay (sampled up to June 30, 2021). A unique set of external references (n=7,992) were obtained by including all the non-Brazilian and non-Paraguayan sequences from the global and South American NextStrain builds (date of access; June 30^th^, 2021). These global references were further supplemented by the inclusion of a random sample of P.1 and P.2 lineages from around the world. This approach was taken to ensure we captured sufficient global sequence diversity while at the same time enriching our data set with South American sequences with whom Brazil and Paraguay share borders. Additionally, during our down sampling strategy, we took into account that the different sequencing efforts employed by different Brazilian states resulted in differing numbers of available genomes for each state/region. For this reason, we selected reference sequences considering both the number of reported cases for each state and the number of available genome sequences, aiming at building a more representative data set (**Fig. S3)**.

### Phylogenetic analysis

Sequences were aligned using MAFFT^44^ and submitted to IQ-TREE2 for maximum likelihood (ML) phylogenetic analysis^46^ employing the general time reversible (GTR) model of nucleotide substitution and a proportion of invariable sites (+I) as selected by the ModelFinder application. Branch support was assessed using the approximate likelihood-ratio test based on the bootstrap and the Shimodaira–Hasegawa-like procedure (SH-aLRT) with 1,000 replicates.

The raw ML tree topology was then used to estimate the number of viral transmission events between various Brazilian regions and the rest of the world. TreeTime^47^ was used to transform this ML tree topology into a dated tree using a constant mean rate of 8.0 × 10^−4^ nucleotide substitutions per site per year, after the exclusion of outlier sequences. A migration model was fitted to the resulting time-scaled phylogenetic tree in TreeTime, mapping country and regional locations to tips and internal nodes^47^. Using the resulting annotated tree topology we were able to count the number of transitions (i.e. virus importations and exportations) within different Brazilian regions and between Brazil and the rest of the world. Importantly, this analysis was not dependent on a monophyletic clustering of SARS-CoV-2 in Brazil. To provide a measure of confidence in the time and source of viral transitions, we performed the discrete ancestral state reconstruction on 10 bootstrap replicate trees (**Fig. S4-S6**).

### Lineage classification

We used the dynamic lineage classification as specified in the Phylogenetic Assignment of Named Global Outbreak LINeages (Pangolin version 3.1.7) protocol^48^. This was aimed at identifying the most epidemiologically important lineages of SARS-CoV-2 circulating within South America (Brazil and Paraguay). Both VOCs and VUMs were designated based on the World Health Organization framework as of July, 2021.

### Phylogeographic reconstruction

VOCs and VUMs that emerged in Brazil (i.e. P.1 and P.2) were identified as monophyletic groups on the time-resolved phylogenetic trees (**Fig.2 and Fig. S7**). Genome sequences from these lineages were then extracted to infer continuous phylogeography histories using the Markov chain Monte Carlo (MCMC) method in BEAST v1.10.4^49^, employing the HKY + Γ_nucleotide substitution model and a strict molecular clock in BEAST v.1.10.4 that assumes constant evolutionary rates throughout the phylogeny.

Due to the huge size of the largest clusters we down-sampled them to <600 taxa per clade to infer phylogeographies within a manageable time frame. Accordingly, we retained the ten earliest sequences from each unique sampling location within Brazil and a random distribution over the remaining samples from each location (n=531 for P.1 and n=428 for P.2). Briefly, sequences from the subsampled cluster were aligned using MAFFT and preliminary ML trees were inferred in IQ-TREE2 as described above. Prior to phylogeographic analysis, each lineage was also assessed for molecular clock signal using the root-to-tip regression method available in TempEst v1.5.3^45^ following the removal of potential outliers that may violate the molecular clock assumption. We accepted temporal structure when the correlation coefficient was > 0.2^50^. Linear regression of root-to-tip genetic distances against sampling dates indicated that the SARS-CoV-2 sequences from Gamma and Zeta evolved in a relatively clock-like manner (r^2^= 0.5; coefficient correlation= 0.45 for Gamma and r^2^= 0.5; coefficient correlation= 0.40 for Zeta) (**Fig. S7**).

We modelled the phylogenetic diffusion and spread of these lineages within Brazil by analysing localized transmission (between Brazilian regions) using a flexible relaxed random walk (RRW) diffusion model^51,52^ that accommodates branch-specific variation in rates of dispersal with a Cauchy distribution and a jitter window site of 0.01^52^. The choice of Cauchy distribution was based on recent studies demonstrating that it is successful for this kind of analyses based on genomes of SARS-CoV-2 and its variants^4,50,54-56^. For each sequence, latitude and longitude coordinates were attributed. MCMC analyses were set up in BEAST v1.10.4, running in duplicate for 100 million interactions and sampling every 10,000 steps in the chain.

Convergence for each run was assessed in Tracer v1.7.1 (ESS for all relevant model parameters >200). Maximum clade credibility trees for each run were summarized using TreeAnnotator after discarding the initial 10% as burn-in. While the sampling is relatively homogeneous among sampled locations, the phylogeographic reconstruction will of course remain sampling-dependent. Hence, differences in sampling effort may have impacted the estimated transition frequencies between locations. Despite this caveat, the phylogeographic analysis still provides important information on the history and dynamics of dispersal of the lineages sampled, which can in turn provide insights into the connectivity of locations along the transmission network. Finally, we used the R package “seraphim”^55^ to extract and map spatiotemporal information embedded in the posterior trees. Note that a transmission link on the phylogeographic map can denote one or more transmission events depending on the phylogeographic inference.

### SARS-CoV-2 time series from Brazil

Data from weekly notified and laboratory confirmed cases of infection by SARS-CoV-2 in Brazil, were supplied by the Brazilian Ministry of Health, as made available by the COVIDA network at https://github.com/wcota/covid19br. For convenience, the geographical locations were aggregated by Brazilian macro regions: North, Northeast, Southeast, South, and Midwest.

## Data Availability

All sequences that were generated and used in the present study are listed in table S1-S3 (accessible on the GitHub repository) along with their GISAID sequence IDs, dates of sampling, the originating and submitting laboratories and main authors. All input files (e.g. alignments or XML files), all resulting output files and scripts used in the study are shared publicly on GitHub (https://github.com/genomicsurveillance/Genomic_epidemiology_reveals_how_restriction_measures_shaped_the_SARS-CoV-2_epidemic_in_Brazil).

## Acknowledgments

We thank all those who have kindly deposited and shared genome data on GISAID (Table S3). The authors acknowledge the technical support of Luciana de Araujo Pimenta, Luiz Aurelio de Campos Crispin, Gabriela Mauric Frossard Ribeiro, Glaucia Maria Rodrigues Borges, Mariane Evaristo e Josiane Serrano Borges from the National Network for Pandemic Alert of SARS-CoV-2 and the contribution of all employees of General Coordination of Public Health Laboratories and professionals of Public Health Laboratories of Brazil and Network for Pandemic Alert of Emerging SARS-CoV-2 Variants for their contribution towards the sequencing effort and for their commitment and work during the fight of the COVID-19 pandemic.

## Author contributions

<u>Molecular screening and produced SARS-CoV-2 genomic data</u>: MG, SNS, VLV, ESR, EVS, FA, JX, HF, TERA, LPOL, AJM, CRSB, ECM, JSTB, DBM, RAB, RLRCC, PDSCM, JPK, BS, RPS, VVC, ST, VBN, LROS, MKAG, JQL, AAR, NRG, LTW, LBS, RSF, MPFP, MJO, SGF, SV, CA, GPMG, WSF, ARMC, JV, SMVLO, CCMG, MCDR, GNB, MPG, NQM, INR, SR, GMC, AHDC, CZ, CDS, PAA, FASC, MDP, JCCL, ECM, CAB, LS, MMM, RMTG, JASN, MLN, HF, LLC, RTC, RMN; <u>Collected samples and curated metadata</u>: CF, CRLP, CFRF, WN, RFCS, MA, CCAM, RH, SCS, MCE, SK, LCJA, DTC, JASN, HF, LLC, FP, AL, FCMI, GCP, CV, GMES, ECO, LD, JC; Analysed the data: MG, VF, EW, HT, JSLP, EJS, RB, JL; <u>Helped with study design and data interpretation</u>: MG, VF, EW, HT, EH, JL, TdO, SCS, MCE, SK, LCJA, DTC; <u>Wrote the initial manuscript, which was reviewed by all authors</u>: MG, SNS, SCS, MCE, SK, LCJA, DTC.

## ADDITIONAL INFORMATION

**Figure S1. Sequencing statistics and lineage assignment of the SARS-CoV-2 genomes generated in this study**. (A) Genome coverage plotted against RT-qPCR cycle threshold value. Colours represent genomes obtained from Brazil (green) and Paraguay (orange), respectively. (B) Number of genomes obtained in this study from Brazil and Paraguay. Colours represent different lineages.

**Figure S2. Fully annotated Brazilian SARS-CoV-2 time tree**. Time resolved maximum likelihood phylogeny containing 17,135 high quality Brazilian SARS-CoV-2 near-full-genome sequences (n=3,866 generated in this study) analysed against a backdrop of global reference sequences. Variants of interest (VUM) and concern (VOC) are highlighted.

**Figure S3**. Number of total cases plotted against the number of sequences available for each location Brazilian state/region.

**Figure S4. Sensitivity of the viral introduction analysis to geographic sampling biases. A, B)** Sensitivity analysis, by performing the discrete ancestral state reconstruction on 10 bootstrap replicate trees, showing how the proportion of international imports and exports into and from Brazil from external locations did not vary in the time and source of viral transitions. **C-K)** Equivalent sensitivity analysis showing how the number of viral exchanges within Brazilian regions by counting the state changes from the root to the tips did not vary in the time and source of viral transitions.

**Figure S5. Sensitivity of the Gamma viral introduction analysis to geographic sampling biases. A, B)** Sensitivity analysis, by performing the discrete ancestral state reconstruction on 10 bootstrap replicate trees, showing how the proportion of international imports and exports of Gamma variant into and from Brazil from external locations did not vary in the time and source of viral transitions. **C-K)** Equivalent sensitivity analysis showing how the number of exchanges of the Gamma variant between Brazilian regions did not vary in the time and source of viral transitions.

**Figure S6. Sensitivity of the Zeta viral introduction analysis to geographic sampling biases. A, B)** Sensitivity analysis, by performing the discrete ancestral state reconstruction on 10 bootstrap replicate trees, showing how the proportion of international imports and exports of Zeta variant into and from Brazil from external locations did not vary in the time and source of viral transitions. **C-K)** Equivalent sensitivity analysis showing how the number of exchanges of the Zeta variant between Brazilian regions did not vary in the time and source of viral transitions.

**Figure S7. Temporal Signal. (**A) Root-to-tip regression plots for Brazilian lineages of concern and of interest (P.1 – Gamma and P.2-Zeta).

**Supplementary Table 1**. Information on the n=3803 SARS-CoV-2 Brazilian samples sequenced as part of this study.

**Supplementary Table 2**. Information on the n=63 SARS-CoV-2 from Paraguay samples sequenced as part of this study.

**Supplementary Table 3**. GISAID acknowledgment table.

